# Assessment of awareness and attitudes of pregnant women toward labor epidural analgesia and factors influencing the decision to opt for it: A cross-sectional study

**DOI:** 10.1101/2025.01.01.25319869

**Authors:** Adugna A. Kassa, Wossenyeleh A. Sahle, Tsehaynew Desalew, Zufan Lakew, Tigist Behabtu

## Abstract

**Background:** Epidural analgesia is one of the most effective forms of pain relief during various stages of labor. Despite its many advantages, the technique is not routinely practiced in most centers in developing countries. Awareness and attitudes toward it, as well as the reasons for its limited utilization, have not been explored in our setting.

Therefore, the aim of this study was to assess the awareness and attitudes of pregnant women toward labor epidurals and to assess the factors influencing their decision to receive labor epidural analgesia (LEA) during labor and delivery.

**Methods:** A cross-sectional study was conducted from June to August 2024 at Hemen Medical Center. A structured questionnaire was used to collect the data. The collected data were analysed via SPSS version 26. Descriptive statistics were used to summarize the descriptive results, and logistic regression was employed to identify factors associated with the decision to receive labor epidural analgesia during delivery. A p value of < 0.05 was used to determine statistical significance.

**Results:** Two hundred thirty pregnant women, with a mean age of 30 ± 4.3 years, were included in the analysis. Approximately 78% of the participants had heard of labor epidural analgesia (LEA), although only 6.5% reported healthcare providers as their primary source of information. Approximately 47% believed that LEA was effective for pain relief during labor. However, only 14.3% expressed a willingness to consider using LEA during delivery. Education level, family income, perceived risk to the Fetus, and perceived effectiveness in relieving pain were identified as factors influencing the decision to consider LEA during delivery.

**Conclusion:** Despite relatively high basic awareness of LEA, many participants remain hesitant to utilize it during labor and delivery. Comprehensive health education and counselling during antenatal care are essential to address misconceptions and promote informed decision-making regarding LEA.

## Introduction

### Background

The majority of women look forward to childbirth as a joyful experience. The worst pain a woman has ever felt nonetheless goes along with it (1). Analgesia during labor and delivery has been approved by the American College of Obstetricians and Gynecologists and the American Society of Anesthesiologists. A maternal request for pain relief during labor has been identified as a sufficient justification (1).

In high-income countries, painless labor is almost universal. However, pain treatment during delivery is still a far-off dream in low-income countries where women are disproportionately burdened by high pregnancy rates and shorter interpregnancy intervals(2).

Epidural analgesia is one of the most effective forms of pain relief during different stages of labor and may allow parturient to rest and relax, facilitating their cooperation during labor and delivery. Despite its benefits, some pregnant women may have reservations about receiving an epidural surgery because of concerns about potential risks and side effects (3).

Therefore, this study aimed to assess the awareness and attitudes of pregnant women towards labor epidural analgesia (LEA) and assess the factors influencing their decision to consider LEA during delivery.

### Statement of the problem

Labor pain is one of the most intense and challenging experiences faced by women during childbirth, necessitating effective pain management for maternal comfort and positive delivery outcomes(3). Epidural analgesia is considered the gold standard for labor pain relief because of its effectiveness and high maternal satisfaction(1). Despite these benefits, the utilization of labor epidural analgesia (LEA) remains low in many developing regions (4,5), including Ethiopia(6). This underutilization stems from a lack of awareness, misconceptions, and fears regarding the procedure and its potential side effects. Cultural beliefs and misinformation further exacerbate negative attitudes toward LEA.

There are no published studies in Ethiopia investigating the awareness and attitudes of pregnant women toward LEA or assessing the factors contributing to its poor utilization.

Understanding these factors is crucial for developing educational interventions that could enhance the acceptance and utilization of LEA.

### Significance of the study

This study aims to fill this critical gap by assessing the awareness and attitudes of pregnant mothers toward labor epidural analgesia and assessing the factors influencing their decision-making through a cross-sectional approach. The insights gained will be helpful in developing targeted educational interventions to address misconceptions and fears and integrated into routine antenatal care to ensure that pregnant women receive accurate and comprehensive information about LEA.

The findings will inform training programs for healthcare providers, equipping them with the ability to counsel expectant mothers effectively. This can create a more supportive environment, enhancing the overall childbirth experience. By contributing data from Ethiopia, this study contributes to the global understanding of how cultural, social, and educational factors influence LEA acceptance, aiding in the design of culturally sensitive educational approaches.

Additionally, increasing awareness and positive attitudes toward LEA can improve maternal and neonatal health outcomes by reducing labor-related stress and anxiety. This study’s findings will promote effective pain management during labor, thus enhancing maternal and neonatal health in Ethiopia.

## Methods

### General objectives

This study aimed to assess the level of awareness and attitudes of pregnant mothers toward labor epidural analgesia and to identify factors affecting their decision to consider LEA during delivery at Hemen Medical Center, Addis Ababa, Ethiopia.

### Specific Objectives

- To determine the level of awareness of labor epidural analgesia among pregnant mothers
- To evaluate the attitudes of pregnant mothers toward labor epidural analgesia.
- To identify the factors that influence the decision of pregnant mothers to consider labor epidural analgesia during delivery.

### Study area

This study was conducted at HMC. Hemen Medical Center is a private maternal and child care center located in Addis Ababa, Ethiopia. The HMC has been one of the pioneer hospitals in introducing and successfully delivering labor epidural analgesia in the private sector since 2012. On average, approximately 25 labor epidurals are performed each month.

### Study Design and Period

An institution-based cross-sectional study was conducted from June 03–August 30, 2024.

### Study Population

All mothers in their 3^rd^ trimester of pregnancy (>28 weeks of pregnancy) attended ANC follow-up at HMC and planned to have a vaginal delivery.

The inclusion criteria for pregnant women were as follows: were aged >18 years, were in their 3^rd^ trimester of pregnancy, planned to have a vaginal delivery, were willing to participate in the study and provided informed consent.

### Exclusion criteria

Patients with a previous history of epidural analgesics and pregnant women with a diagnosed fetal anomaly during the current pregnancy were excluded, as psychological and emotional distress may influence their attitudes and decision-making differently from those of the general population.

### Sample size determination

Sample size was determined via the single proportion formula and finite population correction formula by assuming P = 0.5, since no similar studies have been conducted in Ethiopia, and a 5% margin of error at the 95% confidence interval was calculated via the following formula:

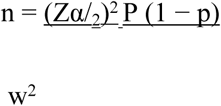

where n = sample size, z= 1.96, p= 0.5, w= 0.05, CI= 95% & ἀ= 5%.

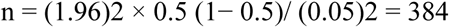

The final sample size, nf = n/(1+n/N), where N= the estimated number of 3^rd^ trimester pregnant women expected to visit HMC for ANC follow-up within the study period, 2 months, is approximately 880.

Therefore, the final sample size is nf = 384/ (1 + 384/880) = **267**. A total sample of 267 3^rd^ trimester pregnant women were included in the study within a 2-month period.

### Sampling Technique

A systematic sampling technique was used. During the data collection period, 880 third-trimester pregnant women were expected to visit the ANC at HMC within three months. Therefore, one out of every three eligible subjects were selected from the ANC appointment lists.

### Dependent variables

Awareness and attitudes of mothers toward labor epidural analgesia and the decision to consider LEA during delivery.

### Independent variables

Maternal age, parity, educational level, income, awareness of epidural analgesia, attitude toward LEA, source of information about labor epidural, and previous childbirth experience.

### Data collection process and technique

Data were collected via a structured, self-administered questionnaire distributed to pregnant mothers during their antenatal visits at the hospital. The questionnaire assessed their level of awareness and attitudes toward labor epidural analgesia, whether they planned to opt for it during delivery, and the factors influencing their decision to choose LEA.

### Data quality assurance

To ensure the quality of the data, training on the objectives and relevance of the study and brief orientations on the assessment tools were provided for the data collectors. After data collection, each questionnaire was checked by the investigator for completeness.

### Data analysis

Completed data were analysed via IBM SPSS Statistics for Windows, version 26 (IBM Corp., Armonk, NY, USA). Descriptive statistics, such as frequencies and percentages, were used to summarize the questionnaire responses. Binary logistic regression was performed to identify the factors influencing pregnant mothers’ decisions regarding LEA use.

### Ethical considerations

The study protocol was reviewed and approved by the Ethiopian Association of Anaesthetists Institutional Research Ethics Review Committee (EAA IRERC) (Protocol Number: EAA/16/30895/005). Data were collected using self-administered questionnaires from pregnant women in their third trimester who were attending antenatal care (ANC) follow-up at Hemen Medical Center between June 3, 2024, and August 30, 2024. Participants who were willing to take part in the study confirmed their consent by signing the information page of the provided questionnaire. They were assured of confidentiality and informed that their participation was entirely voluntary. To maintain privacy, the completed questionnaires were securely stored in a locked location.

## Results

### Demographic characteristics of the participants

A total of 267 pregnant women attending antenatal care (ANC) follow-up at Hemen Medical Centre participated in the study. Of these, 37 either did not return the questionnaire or submitted it incomplete. Therefore, 230 participants were included in the analysis. The mean age of the participants was 29.9 <u>±</u> 4.3 years, and approximately 70% of the participants were in the 25--34 years age group. The majority of the respondents (66.6%) were degree holders or higher, and approximately 75% were employed in various occupations. The monthly family income varied, with approximately 20% earning less than 10,000 ETB, 11% reporting an income above 50,000 ETB, and 1 USD earning approximately 100ETB. In terms of obstetric history, the mean gestational age of the participants was 37.8 ± 1.8 weeks, and approximately half of the participants were nulliparous (Table 1).

**Table 1:** Sociodemographic and obstetric characteristics of third trimester pregnant women attending ANC follow-up at HMC, Ethiopia

| Variables | Category | Frequency(n) | Percentage |
| --- | --- | --- | --- |
| Maternal age group | 18-25 years old | 22 | 13.2% |
|  | 26-34 years old | 117 | 70% |
|  | ≥35 years old | 28 | 16.8% |
| Level of Education | Primary School | 9 | 4% |
|  | Secondary School | 27 | 11.7% |
|  | Diploma | 41 | 17.8% |
|  | Degree | 100 | 43.5% |
|  | Masters and above | 53 | 23% |
| Occupation | Unemployed | 7 | 3% |
|  | Stay at home mother | 51 | 22.2% |
|  | Government Employee | 61 | 26.5% |
|  | Private Employee | 70 | 30.4% |
|  | Private business owner | 41 | 17.8% |
| Monthly family income | <10, 000 ETB | 46 | 20% |
| | 10,000 – 25,000 ETB<br>(\$100 - \$250) | 84 | 36.5% |
| | 25,000 – 50,000 ETB<br>(\$250 - \$500) | 74 | 32.2% |
| | >50, 000 ETB<br>(>\$500) | 26 | 11.3% |
| Parity | Nulliparous | 112 | 48.7% |
|  | Primiparous | 66 | 28.7% |
|  | Multiparous | 52 | 22.6% |

### Awareness of labor epidural analgesia

Among the total participants, 78% reported having heard of labor epidural analgesia (LEA). The primary sources of information were previous users (27%), followed by family and friends (24%) and the internet (19.6%). Only 6.5% of the respondents cited healthcare providers as their primary source, underscoring the need for direct communication to enhance awareness. Additionally, only 34% of the participants provided explanations from health care providers about LEA during their ANC follow-up.

Moreover, a significant portion of the participants (21.7%) had not previously heard of LEA.

### Attitudes Toward Labor Epidural Analgesia

Regarding attitudes, 47% of women believed that LEA was effective for pain relief during labor. However, only 14.3% expressed a willingness to consider using LEA during delivery, while the rest did not have a plan at all (47.8%) or were undecided to take LEA (37.8%). The main concerns among those unwilling or hesitant to use LEA included fear of side effects for the mother (20%), concerns for the baby’s safety (11%), interference with the natural labor process (11%), and additional cost (20%).

### Factors influencing decision-making regarding LEA during laboring

Bivariate logistic regression analysis revealed that education level, occupation, family monthly income, perception of LEA causing significant problems for the mother and Fetus, and awareness of its pain-relief effectiveness were factors associated with the decision to receive LEA, with a p value < 0.2. These variables were included in the multivariable logistic regression analysis.

The multivariable logistic regression analysis revealed that education level, family monthly income, the perception that LEA could cause significant problems for the Fetus, and the perception of LEA effectiveness in relieving pain were significantly associated with the decision to receive LEA during labor and delivery, with a p value < 0.05 (Table 2).

**Table 2:** Factors influencing the decision to receive LEA during labor of participants at HMC, Ethiopia.

| Variables |  | Decision to take LEA |  | COR (95% CI) | AOR (95% CI) | P value |
| --- | --- | --- | --- | --- | --- | --- |
|  |  | Yes | No/Not sure |  |  |  |
| Education | Diploma and Lower | 3 (4.4%) | 66 (95.6%) | 1 | 1 | **0.021\*** |
|  | Degree and above | 30 (18.6%) | 131 (81.4%) | 5.01 (1.48, 17.1) | 1.9 (1.28, 6.78) |  |
| Occupation | Unemployed | 5 (8.6%) | 53 (91.4%) | 1 | 1 | 0.402 |
|  | Employed | 28 (16.3%) | 144 (83.7%) | 2.1 (1.06, 5.6) | 1.6 (0.53, 4.8) |  |
| Income | <25,000 ETB | 12 (9%) | 109 (91%) | 0.31 (0.3, 0.79) | 0.28 (0.11, 0.69) | **0.006\*** |
|  | ≥25,000 ETB | 21 (19.3%) | 88 (80.7%) | 1 | 1 |  |
| Perceived problem on mother | No | 17 (22.6%) | 58 (77.4%) | 1 | 1 | 0.65 |
|  | Yes/Not sure | 16 (10.3%) | 139 (89.7%) | 0.4 (0.19 – 0.83) | 0.82 (0.34, 1.96) |  |
| Perceived problem on Fetus | No | 23 (29.1%) | 56 (70.9%) | 1 | 1 | <**0.001\*** |
|  | Yes/Not sure | 10 (6.6%) | 141 (93.4%) | 0.17 (0.08 – 0.39) | 0.18 (0.07, 0.45) |  |
| Pain relief perception | Yes | 27 (25%) | 81 (75%) | 1 | 1 | **0.001\*** |
|  | No/not sure | 6 (4.9%) | 116 (95.1%) | 0.16 (0.06, 0.4) | 0.18 (0.06, 0.49) |  |

Note: LEA = labor epidural analgesia, COR = crude odds ratio, AOR = adjusted odds ratio, CI =
confidence interval
\*Statistically significant association at $P < 0.05$

## Discussion

### Awareness of Labor Epidural Analgesia

This study revealed that 78% of the participants had prior awareness of labor epidural analgesia (LEA). Among those who were aware, only 6.5% identified healthcare providers as their primary source of information. Furthermore, only 14.3% of the participants expressed a willingness to receive LEA during delivery. The main reasons for hesitancy or unwillingness to accept LEA included concerns about potential side effects for the mother and baby, perceived interference with the labor process, and the additional cost associated with the technique.

The relatively high level of awareness observed in this study may be attributed to nearly all participants (98%) being from urban areas, specifically Addis Ababa, the capital city of Ethiopia. Urban residency likely increases exposure to advanced obstetric practices, either directly or indirectly (7).

Our findings are consistent with studies conducted in urban settings. For example, a study among women residing in Riyadh reported that most participants demonstrated good knowledge of epidural analgesia for labor pain relief (8). Similarly, another study conducted in the same region also documented adequate knowledge among participants regarding epidural analgesia (9).

In contrast, numerous studies conducted in developing countries have reported low levels of awareness of labor analgesia. For example, two Ethiopian studies highlighted that the overall awareness of labor analgesia among pregnant women was low (7,10). A study conducted in Nigeria also reported low awareness levels of labor epidural analgesia among its participants (11). Similarly, studies from India identified a lack of awareness about labor epidural analgesia in the majority of their study populations (3,12).

### Attitude toward Labor Epidural Analgesia

Our study revealed that only 14.3% of participants were willing to consider labor epidural analgesia (LEA) during delivery, despite 47% acknowledging its effectiveness in relieving labor pain. The primary reasons for their reluctance included concerns about potential side effects for both the mother and the Fetus. However, these perceptions are largely unfounded, as recent studies have demonstrated the safety of LEA for both maternal and fetal outcomes (13–15).

### Factors influencing the decision to perform LEA during labor

Our study revealed that educational level, monthly family income, concern regarding the potential adverse effects of LEA on fetal well-being, and perceived efficacy of LEA in pain relief were factors influencing the decision to perform LEA during labor and delivery.

Compared with those with a diploma or lower educational attainment, those with a degree or higher education are twice as likely to opt for LEA (P = 0.021, AOR: 1.9). In line with our findings, a study conducted in Ethiopia reported that a higher education level is associated with a positive attitude toward labor analgesia(10). Multiple other studies conducted in different parts of the world have also identified the level of education as an important factor influencing pregnant women’s decision to request LEA (8,16–18). This association may be explained by the fact that individuals with higher education typically have greater access to reliable health information and are more likely to engage in effective communication with healthcare providers. These factors contribute to an enhanced understanding and acceptance of LEA as an evidence-based option for pain management during labor (3,17,18).

In this study, monthly family income was identified as a factor influencing women’s decision to utilize labor epidural analgesia (LEA). According to our findings, women with a monthly family income exceeding 25,000 ETB ($250) were 3.5 times more likely to seek LEA during labor and delivery than were those with a lower income (P = 0.006, AOR: 0.28). Multiple studies have similarly reported that income is significantly associated with the decision to opt for LEA (8,17,19). A possible explanation for this finding is that LEA adds to the overall cost of care. The additional expense associated with the technique was frequently mentioned by our study participants as a reason for their unwillingness to choose LEA.

Perceived concerns about potential harm to the Fetus emerged as a significant factor influencing the willingness to accept LEA in our study. The participants who believed that LEA caused significant harm to their baby were more than five times more likely to refuse LEA during labor and delivery (P< 0.001, AOR = 0.18). Consistent with our findings, previous studies have reported similar observations(3,20). However, substantial evidence indicates that LEA does not pose any risk to the Fetus (14,15,21); in contrast, it may confer benefits, such as improved oxygenation and higher APGAR scores at delivery (13,22). These results highlight how misconceptions and a lack of health education can negatively influence decisions regarding quality healthcare. The evidence suggests that health education can play a critical role in improving women’s decisions to opt for LEA during labor and delivery (23,24).

Our study also revealed that pregnant women who believe that LEA is effective in preventing labor pain are more than six times more likely to choose LEA during delivery (P = 0.001, AOR = 0.18). Similarly, other studies have reported that when pregnant women perceive LEA as effective in relieving labor pain, the likelihood of choosing it significantly increases (12,23–25). These findings underscore the importance of educating women about the effectiveness of LEA in managing labor pain during antenatal care follow-up visits.

## Limitations

Our study has some limitations. First, we were unable to assess the impact of previous epidural experience on maternal decisions regarding labor epidural analgesia (LEA), as LEA is a relatively new practice in our country and few women have prior exposure to it. Second, the study was conducted at a single medical center, which may limit the generalizability of the findings to other regions in Ethiopia, as the sample may not fully represent the broader population. Lastly, the cross-sectional design of the study precluded us from tracking participants to determine how many ultimately opted for LEA during delivery.

## Conclusions

This study highlights that while basic awareness of LEA is relatively high, many participants remain hesitant to utilize it during labor and delivery. This reluctance appears to stem from inadequate explanations provided by healthcare professionals, potentially leading to negative perceptions. The factors significantly associated with participants’ decisions to seek LEA included educational level, income, perceived risks to the Fetus, and perceptions of pain relief effectiveness. These findings underscore the need for comprehensive health education and counselling during antenatal care to address misconceptions and promote informed decision-making regarding LEA.

## Data Availability

Data can be made available to researchers upon request by contacting the authors.

## List of abbreviations

ANC: Antenatal Care
AOR: adjusted odds ratio
ASA: American Society of Anesthesiologists
CI: confidence interval
COR: Crude odds ratio
EAA IRERC: Ethiopian Association of Anaesthetists Institutional Research Ethics Review Committee
ETB: Ethiopian Birr
HMC: Hemen Medical Center
LEA: labor epidural analgesia
USD: United States Dollar

## Acknowledgements

We extend our gratitude to the Hemen Maternal and Child Health Centre and its staff for their invaluable support in facilitating this study. We also deeply appreciate the participating women for sharing their experiences, which were instrumental to this research.

